# Temporal Learning with Dynamic Range (TLDR) for Modeling Recurrent Exposure and Treatment Outcomes

**DOI:** 10.1101/2025.03.19.25324272

**Authors:** Jingya Cheng, Jonas Hügel, Jiazi Tian, Alaleh Azhir, Shawn N. Murphy, Jeffrey G. Klann, Hossein Estiri

**Author notes:** Equal contribution (co–first authors) (Jingya Cheng, Jonas Hügel). Co–senior authors (Jeffrey G. Klann, Hossein Estiri). Corresponding, 399 Revolution Drive, Suite 790, Somerville, MA, 02145, USA.

## Abstract

**Background:** The temporal sequence of clinical events is crucial in outcomes research, yet standard machine learning (ML) approaches often overlook this aspect in electronic health records (EHRs), limiting predictive accuracy.

**Methods:** We introduce Temporal Learning with Dynamic Range (TLDR), a time-sensitive ML framework, to identify risk factors for post-acute sequelae of SARS-CoV-2 infection (PASC). Using longitudinal EHR data from over 85,000 patients in the Precision PASC Research Cohort (P2RC) from a large integrated academic medical center, we compare TLDR against a conventional atemporal ML model.

**Results:** TLDR demonstrated superior predictive performance, achieving a mean AUROC of 0.791 compared to 0.668 for the benchmark, marking an 18.4% improvement. Additionally, TLDR’s mean PRAUC of 0.590 significantly outperformed the benchmark’s 0.421, a 40.14% increase. The framework exhibited improved generalizability with a lower mean overfitting index (−0.028), highlighting its robustness. Beyond predictive gains, TLDR’s use of time-stamped features enhanced interpretability, offering a more precise characterization of individual patient records.

**Discussion:** TLDR effectively captures exposure–outcome associations and offers flexibility in time-stamping strategies to suit diverse clinical research needs.

**Conclusion:** TLDR provides a simple yet effective approach for integrating dynamic temporal windows into predictive modeling. It is available within the MLHO R package to support further exploration of recurrent treatment and exposure patterns in various clinical settings.

## 1 Introduction

In clinical care, the prognostic value of a patient’s history is not uniform; an event’s timing relative to an outcome is often what determines its importance. While Transformer models solve a similar problem in natural language by using soft, learned attention to dynamically weight tokens based on contextual relevance[1], the irregular, sparse, and asynchronous nature of electronic health records (EHRs) calls for a more structured approach. For clinical sequences, dense softmax attention can be computationally inefficient and lacks interpretability.

We introduce Temporal Learning with Dynamic Range (TLDR), a hard-attention operator specialized for EHR data. TLDR defines fixed temporal windows (‘history’, ‘past’, ‘last’) that act as queries. It aligns clinical events as keys and values and replaces soft, learned weighting with an information-theoretic selection criterion (MSMR) that functions as a form of hard attention. This process yields a compact, interpretable temporal representation that preserves clinically salient signals while discarding noise. By embedding temporal relevance directly into the feature space, TLDR bridges formal temporal reasoning and modern attention-based learning, offering a principled alternative for modeling complex health records.

In temporal outcomes research, particularly in settings involving repeated exposures, the sequential ordering of events holds significant prognostic value. For instance, in a patient with diabetes, symptoms like dizziness during a recent insulin adjustment may offer greater predictive utility for hypoglycemia than a static, atemporal collection of all historical data. While traditional biostatistics employs repeated-measures designs for structured longitudinal data[2, 3, 4, 5, 6], leveraging the temporal information in EHRs is challenging due to issues like documentation lag and asynchronous event recording[7, 8]. Consequently, most standard ML approaches fail to account for this informative temporal structure, limiting their ability to capture underlying clinical dynamics[11]. TLDR is designed to overcome these limitations by effectively leveraging the temporal structure of EHRs in downstream ML applications[10, 7, 12, 13], enhancing predictive accuracy for outcomes such as cancer[14, 15], Alzheimer’s disease[8], and sepsis[16, 17].

Here, we apply TLDR to identify risk factors for post-acute sequelae of SARS-CoV-2 (PASC), a condition where individuals may experience multiple infection episodes, making it an ideal use case for studying recurrent exposures[18, 19, 20]. We compare its performance against traditional atemporal modeling.

## 2 Method

### 2.1 Cohort

The validated Precision PASC Research Cohort (P2RC)[20] from Mass General Brigham (MGB) comprises integrated longitudinal clinical data from over 85,000 patients within 12 months of COVID-19 infection. Data were collected across eight hospitals and community health centers in Massachusetts. Inclusion was restricted to infections between March 6, 2020 and June 7, 2022, with follow-up through June 7, 2023. All SARS-CoV-2 positive patients were included. Patients who developed PASC within 12 months were labeled PASC; others were labeled Non-PASC. Use of data was authorized under MGB IRB protocol #2020P001063 with waiver of consent.

### 2.2 Temporal Learning with Dynamic Ranges (TLDR)

We introduce TLDR, a framework for mining temporal structure in EHRs that operates analogously to attention mechanisms. Whereas Transformers employ *soft attention* to compute weighted averages of all past tokens, TLDR implements a form of *hard attention* by directly selecting a sparse subset of temporally contextualized features, yielding both computational efficiency and interpretability.

#### Key Idea

TLDR replaces dense softmax-based aggregation with discrete, information-theoretic selection of temporal events. Instead of distributing fractional weights across all inputs, it identifies and retains only the most informative ones.

#### Intuition

Clinicians rarely consider every past lab value or diagnosis when assessing a patient; rather, they focus on a handful of temporally salient cues. TLDR mirrors this practice by extracting a compact set of features that maximally preserve predictive information.

### 2.3 Temporal Representation Mining

Let each patient *p* have a sequence of timestamped clinical events 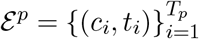, where *c*_*i*_ ∈ 𝒞 denotes a clinical encounter and *t*_*i*_ ∈*T* its time of occurrence. Let *T* = {*t*_1_, *t*_2_, …, *t*_*m*−1_, *t*_outcome_} denote the sequence of time points corresponding to exposures and outcome, where *t*_1_ is the first exposure, *t*_*m*−1_ the final exposure prior to the outcome, and *t*_outcome_ the observed outcome.

We introduce buffer parameters *B* = {*B*_history_, *B*_past_, *B*_last_, *B*_outcome_}, corresponding to the categories *history, past, last*, and *outcome*, respectively. Each buffer *B*_·_specifies a time interval excluded from the boundary of its associated window.

We then categorize each trajectory into three mutually exclusive temporal strata:

- **History** 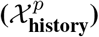: All clinical events occurring prior to the first exposure after accounting for the history buffer:

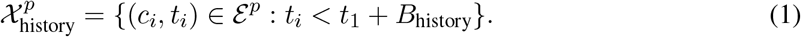
- **Past**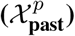: Events between the first exposure and the last exposure before the outcome, excluding buffer regions:

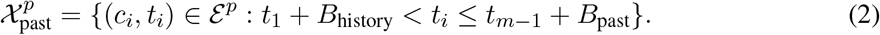
- **Last** 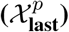: Events in the temporal vicinity of outcome measurement, spanning from the last exposure (adjusted by *B*_past_) through the outcome time (adjusted by *B*_last_ and *B*_outcome_):

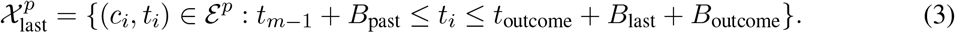

For each concept *c* ∈ 𝒞and temporal stratum *ω* ∈ {history, past, last}, we define a binary incidence variable:

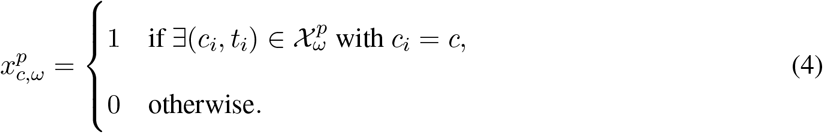

This yields a structured sparse patient-level representation *X*^*p*^ ∈ {0, 1}^|C|×3^,where each clinical encounter is explicitly contextualized within the three temporal strata: *history, past*, and *last*.

### 2.4 Hard Attention via MSMR

To mitigate redundancy and improve discriminative efficiency, TLDR incorporates a discrete attention mechanism based on the *Minimize Sparsity, Maximize Relevance* (MSMR) algorithm. Each candidate feature (*c, ω*), defined by clinical concept *c* ∈ 𝒞 and temporal stratum *ω* ∈ {history, past, last}, is scored by its mutual information with the outcome label *y*:

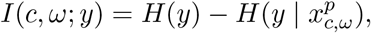

where *H*(·) denotes entropy and 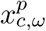 is the binary incidence variable from Eq.4.

MSMR employs a greedy forward-selection strategy. At iteration *r*, given a selected feature set *S*_r−1_, the algorithm adds the feature with maximal conditional mutual information:

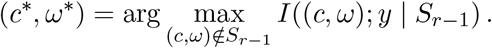

After *R* iterations, the final set *S*_*R*_ defines a sparse representation.

### 2.5 Benchmark (Atemporal Representation)

As a benchmark, we construct an *atemporal* representation that ignores temporal stratification. Each patient trajectory is collapsed into aggregated concept counts:

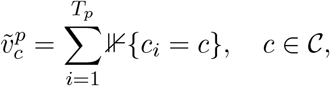

where 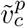 denotes the total number of occurrences of concept *c* in patient *p*’s record.

For each patient *p*, this yields a vector 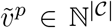, with one dimension per clinical event. Collecting all patients produces the atemporal feature matrix 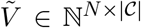, where *N* is the number of patients. Unlike TLDR, which encodes temporally segmented features 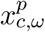 across *history, past*, and *last* windows, the atemporal baseline collapses all events into a single count per concept. This corresponds to the conventional “bag-of-codes” representation that discards temporal information entirely.

### 2.6 Dimensionality Reduction via JMI

Both temporal (TLDR) and atemporal representations are pruned using the *Joint Mutual Information* (JMI) criterion[24], implemented via MLHO’s `MSMR.lite`. JMI balances relevance to the outcome with redundancy among features. For a candidate feature *f*_*k*_, the objective is

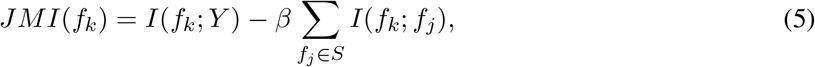

where *S* is the set of already selected features and *β >* 0 controls the relevance–redundancy trade-off. The procedure yields maximally informative, minimally correlated feature subsets, restricted to fewer than 200 features per partition; however, this threshold is a tunable hyperparameter that may be adjusted according to data scale and model capacity.

### 2.7 Experimental Design

The dataset was stratified into 17 non-overlapping subsets. The first 16 subsets contained approximately 5,000 patients each, with the final subset including the remaining patients. On each subset, we trained four classical learning algorithms: Gradient Boosting Machines (GBM), Generalized Linear Models (GLM), L1-regularized logistic regression, and XGBoost—yielding 17 × 4 = 68 predictive models. As deep learning baselines, we implemented BioClinicalBERT[25] and ELECTRA[26] using HuggingFace Transformers (v4.38.2) and PyTorch (v2.1.2). Patient histories were represented as concatenated ICD code sequences, truncated to 128 tokens (BioClinicalBERT) and 256 tokens (ELECTRA).

### 2.8 Evaluation Metrics

Each subset was split into 70% training and 30% testing. Performance was evaluated by mean Area Under the Receiver Operating Characteristic curve (AUROC) and mean Area Under the Precision–Recall curve (AUPRC). Overfitting was assessed by the discrepancy between validation and test AUROC. Statistical significance of TLDR relative to baselines was evaluated via Wilcoxon signed-rank tests with Benjamini–Hochberg multiple-comparison correction.

### 2.9 Schematic Comparison

Figure1 illustrates the conceptual analogy between TLDR and Transformer architectures. Transformers compute dense, soft attention weights across all tokens, yielding contextual embeddings via weighted summation. By contrast, TLDR first partitions events into clinically grounded temporal windows (Section 2.3), then applies MSMR to enforce a hard, binary attention mask, propagating only the most informative features to downstream models.

**Figure 1.**
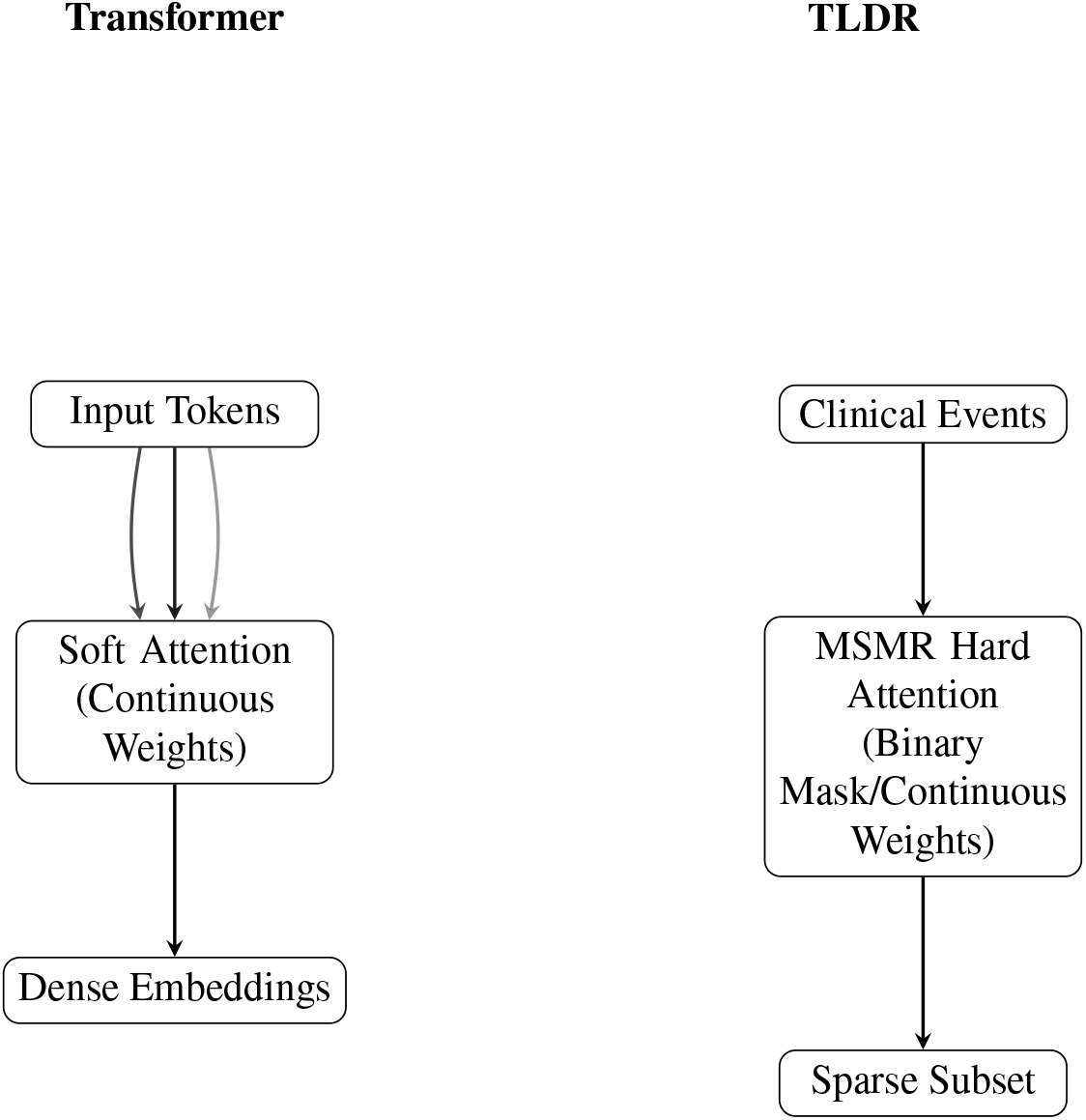
Comparison of Transformer (soft attention) and TLDR (hard attention). Soft attention assigns continuous weights to all inputs, producing dense embeddings. TLDR enforces either binary selection or continuous weights over temporally stratified events, producing sparse yet highly informative feature subsets.

### 2.10 Algorithmic Framework

#### Algorithm 1

TLDR: Temporal Learning with MSMR Hard Attention

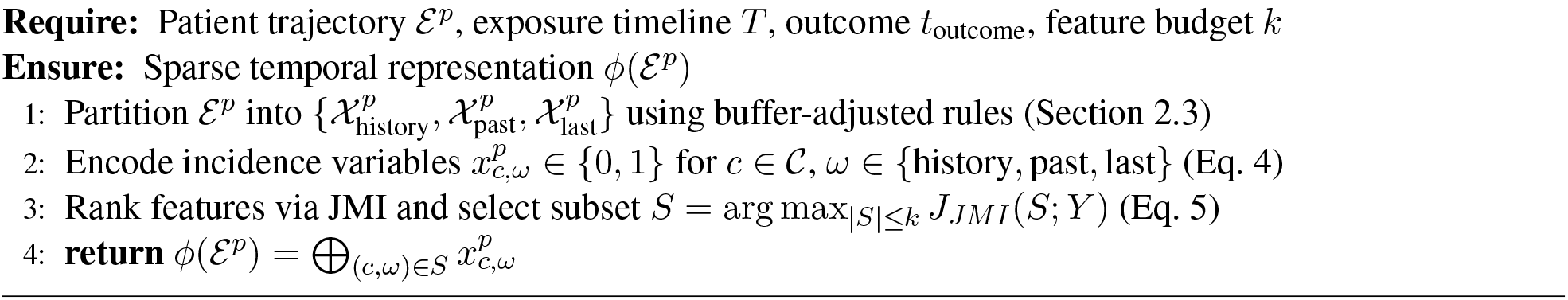

## 3 Results

In the MGB P2RC, which encompasses 85,376 patients, 24,360 individuals developed PASC within 12 months of their infection, while 61,016 did not (Table1). The cohort’s mean age was 53.6 years. Approximately 62.5% of participants were female, 71.4% were White, 10% were Black or African American, and 7.2% were Hispanic. Among those classified as long-haulers, the mean age was 57.1 years. Approximately 64.5% of this subgroup were female, 71.4% were White, 10.4% were Black or African American, and 6.6% were Hispanic. The mean Charlson Comorbidity Index was 2.18 for the overall cohort and 3.06 among the long-haulers.

**Table 1:**
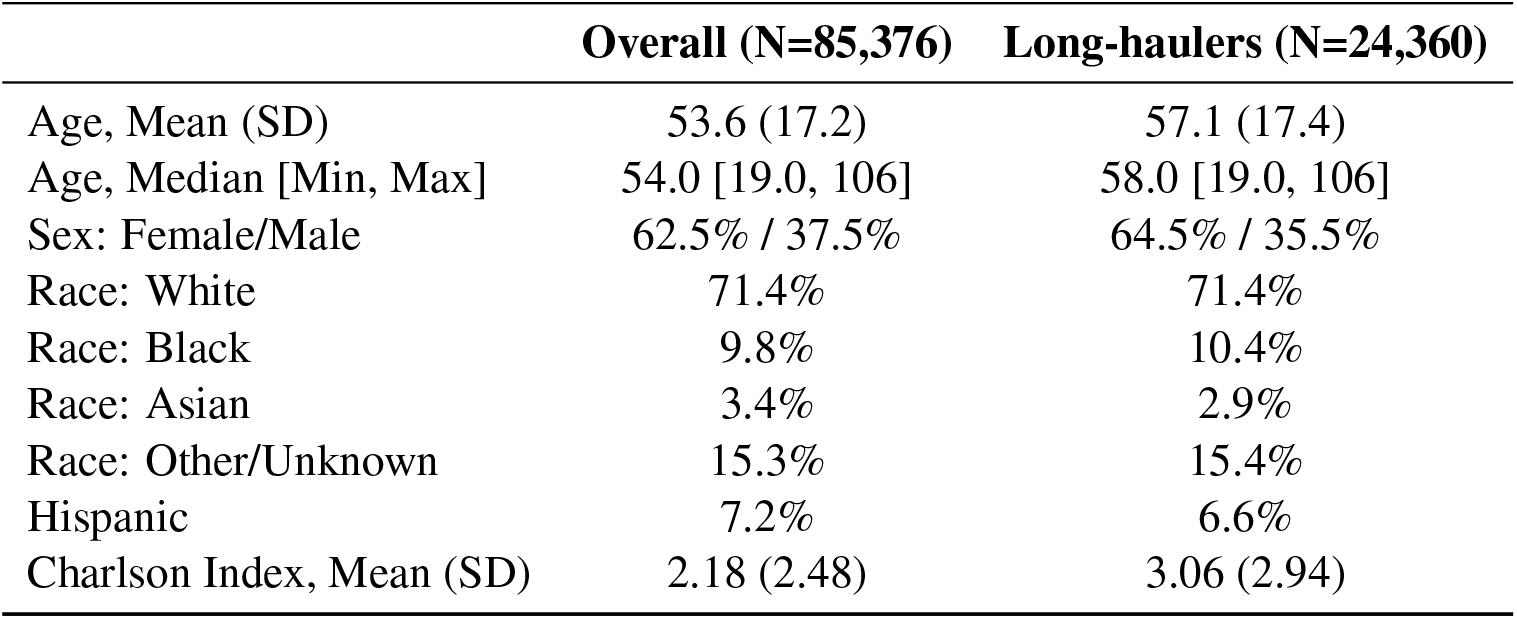
Summary statistics of the study population.

**Table 2:** Model performance metrics for benchmark and TLDR approaches (means across 17 subsets).

|  | AUROC (95% CI) | PRAUC | Overfitting Index |
| --- | --- | --- | --- |
| Benchmark | 0.668 (0.640, 0.668) | 0.421 | -0.001 |
| **TLDR** | **0.791** (0.767, 0.791) | **0.590** | **-0.028** |
| $\Delta$ (TLDR-Benchmark) | +0.123 ($p < 0.001$) | +0.169 ($p < 0.001$) | -0.027 ($p < 0.001$) |

**Table 3:** Model performance by learner (mean AUROC; 95% CI).

| Model | Benchmark | TLDR |
| --- | --- | --- |
| GBM | 0.681 (0.673, 0.689) | **0.810** (0.804, 0.815) |
| GLM | 0.652 (0.644, 0.660) | **0.769** (0.762, 0.776) |
| L1 Logistic | 0.662 (0.655, 0.670) | **0.784** (0.776, 0.792) |
| XGBoost | 0.678 (0.669, 0.670) | **0.803** (0.797, 0.809) |
| Transformer: ELECTRA | 0.699 (0.682, 0.715) |  |
| Transformer: BioClinicalBERT | 0.781 (0.755, 0.807) |  |

### Model outputs

Our analysis utilized 2,336 unique diagnostic features, 1,795 procedural features, 1,240 medication-related features, and 296 laboratory features before dimensionality reduction. The mean AU-ROC for the TLDR approach was 0.791, compared to 0.668 for the benchmark, an 18.4% improvement. The mean PRAUC for TLDR was 0.590, significantly higher than the benchmark’s 0.421, a 40.14% increase. TLDR also exhibited superior generalization with a mean overfitting index of −0.028. Statistical significance was confirmed with Wilcoxon tests.

### Model Benchmark TLDR

TLDR outperformed both transformer-based models, with an average AUC that was 13.0% higher than ELECTRA and 1.41% higher than BioClinicalBERT. Notably, the GBM model of TLDR exhibited an average AUC 15.71% higher than ELECTRA and 3.85% higher than BioClinicalBERT.

### Feature examples

Temporal features create a stronger predictive model by considering the feature only when it is important. *Z79 Long-Term (Current) Drug Therapy [last]* is one such example. Surgical procedures on arteries and veins [past/last] appear near the top of feature importance lists, while the [history] version appears near the bottom.

## 4 Discussion

Our study demonstrates that TLDR offers substantial gains in predictive performance by explicitly encoding temporal context. Conceptually, TLDR can be viewed as a form of hard attention over longitudinal health records. Whereas Transformer architectures compute soft weights over all past tokens, TLDR enforces a structured partition of the clinical timeline into ‘history’, ‘past’, and ‘last’. These partitions act as queries that condition which events (keys/values) are relevant, and MSMR performs information-theoretic feature selection that parallels the weighting step in attention. In effect, TLDR implements a deterministic, domain-aware attention operator that selects features whose presence in specific temporal windows maximally informs the outcome.

This distinction is clinically meaningful. In many EHR settings, the challenge is dealing with irregular, sparse, and asynchronous sequences where event timing carries strong prognostic signals. By partitioning event space into interpretable windows, TLDR surfaces relationships that may be obscured by fully datadriven attention. For example, diagnoses recorded in the ‘last’ window following an exposure often signal acute decompensation, whereas the same diagnoses in ‘history’ carry far less weight. While Transformer models can in principle learn this distinction, they often require substantial data and lose interpretability. TLDR achieves this separation with simple temporal operators and minimal computational overhead.

The empirical comparison underscores this: while BioClinicalBERT benefited from domain-specific pretraining, TLDR outperformed both transformer baselines by embedding temporal relevance directly into the representation. This bridges the gap between symbolic temporal reasoning and neural attention, offering a scalable alternative that maintains interpretability. Beyond its enhanced predictive performance, timestamped features in TLDR provide greater precision in characterizing individual EHRs, thereby improving interpretability. For example, *Long-Term Drug Therapy* documented after the last infection likely signifies ongoing treatment for chronic conditions that may increase PASC risk. Likewise, *Sleep Disorders* and *Abnormalities of Heart Rate* were influential when recorded in historical data, suggesting they may be early indicators of long-COVID risk.

### 4.1 Clinical Implications

From a clinical perspective, TLDR highlights a middle path between atemporal counts and deep sequential models. Atemporal counts (bag-of-codes models) disregard sequence information, while Transformers capture fine-grained dynamics but demand massive pretraining and GPU resources[9, 11]. TLDR offers a pragmatic compromise: capturing coarse temporal context without sacrificing efficiency or interpretability. Recent EHR-specific Transformers such as Med-BERT[27], ClinicalBERT[25], and newer architectures like Foresight[29] or the lung cancer diagnostic Transformer[30] achieve impressive benchmarks but remain opaque and resource-intensive, whereas TLDR yields sparse, auditable representations trainable on modest infrastructure. This design parallels clinical reasoning itself—physicians weigh the recency of diagnoses and interventions differently when assessing prognosis[31, 32]. By encoding this intuition algorithmically, TLDR supports predictive modeling in real-world settings where transparency, reproducibility, and accountability are essential. In particular, TLDR may be attractive for health systems seeking scalable risk prediction without the opacity of black-box embeddings.

#### Limitations and Future Directions

A limitation of our current approach is that the temporal windows are predefined and fixed. The method also relies on clearly defined index dates for exposures and outcomes, which may not be available for all clinical problems. Future work could address these limitations by exploring methods to learn optimal temporal window boundaries directly from the data. Furthermore, the concept could be extended to a “multi-head hard-attention” framework, where different “heads” could attend to different feature types (e.g., diagnoses vs. labs) or different temporal scales. Finally, hybrid models that combine the interpretable, structured features from TLDR with the expressive power of a smaller Transformer could potentially achieve the best of both worlds, balancing performance with clinical utility.

## 5 Conclusion

TLDR offers a straightforward yet effective means of integrating dynamic temporal windows into predictive modeling. Formulated as a temporal hard-attention operator, it provides interpretable, compact representations that improve performance and generalization in EHR tasks with recurrent exposures and outcomes. The method is available within the MLHO R package.

## Supporting information

TLDR examples

## Data Availability

The R package associated with this study can be accessed at the following GitHub repository: https://github.com/clai-group/MLHO.

## Data and Code Availability

Due to patient privacy regulations, the dataset is not publicly available. The R package is available at https://github.com/clai-group/MLHO.

## Ethics Approval

Use of patient data in this study was approved by the Mass General Brigham Institutional Review Board (protocol 2020P001063).

## Acknowledgments

Supported by NIAID R01AI165535. J. Hügel was partially funded by DAAD IFI, BMBF, and DFG (426671079).

## Author Contributions

H.E., J.C., J.G.K., and J.H. conceived, designed, and planned the study. H.E., J.C., and J.G.K. collected and acquired the data. H.E., J.C., and J.G.K. performed data preparation. J.C. and H.E. analyzed the data. H.E., J.C., A.A., S.N.M., J.H., and J.G.K. interpreted the data. H.E., J.C., J.G.K., and J.H. drafted the paper. All authors critically reviewed and revised the final paper. All authors approved the decision to submit for publication.

