## Supplementary material for "Temporal Learning with Dynamic Range (TLDR) for Modeling Recurrent Exposure and Treatment Outcomes": TLDR examples

### Without Outcome date

labelDT:

| Patient ID | Start date |
|---|---|
| 1 | 2020-02-04 |
| 1 | 2020-08-16 |
| 1 | 2021-07-15 |
| 1 | 2022-04-05 |

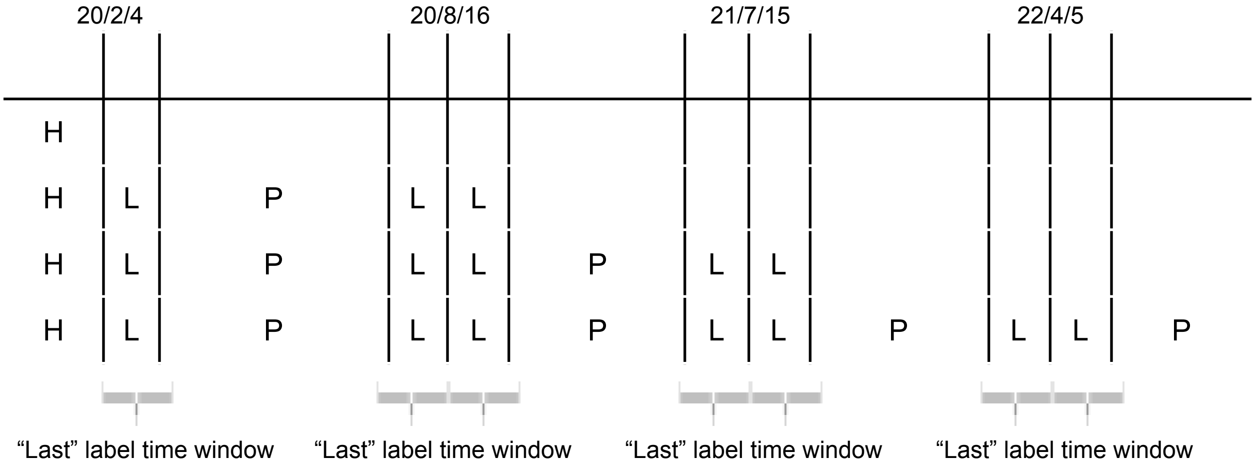


### With Outcome date

#### **No outcome** for all the exposures/treatment

labelDT:

| Patient ID | Start date | Outcome date | label |
|---|---|---|---|
| 1 | 2020-02-04 | 2020-02-04 | 0 |
| 1 | 2020-08-16 | 2020-08-16 | 0 |
| 1 | 2021-07-15 | 2021-07-15 | 0 |
| 1 | 2022-04-05 | 2022-04-05 | 0 |

Number of distinct Incidences by each Label. (Eg. there are 4 distinct incidences for all features that are labeled as “history”. For all features from 20/2/4 to 20/8/16 that are labeled as “past” events, there are 2 distinct incidences)

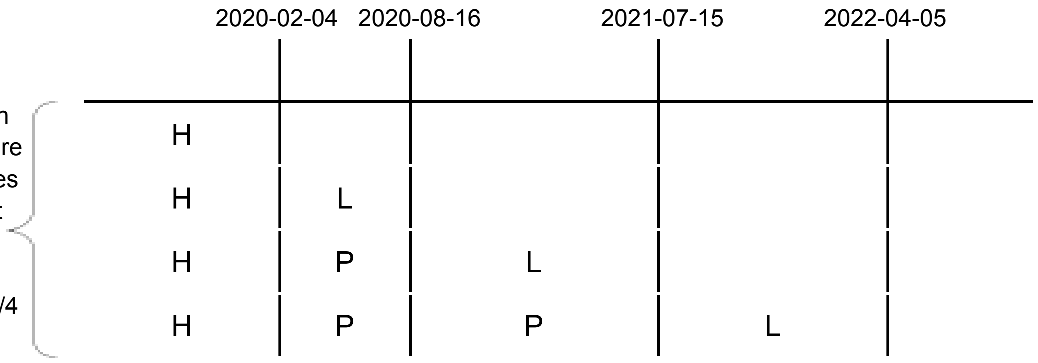


#### With outcome(s)

##### Example 1

| Patient ID | Start date | Outcome date | label |
|---|---|---|---|
| 1 | 2020-02-04 | 2021-03-16 | 1 |

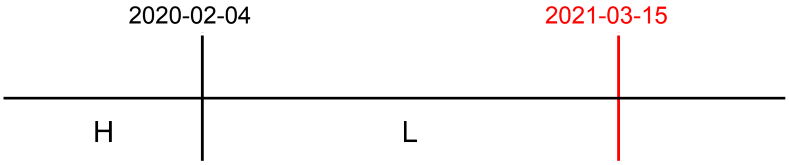


##### Example 2

| Patient ID | Start date | Outcome date | label |
|---|---|---|---|
| 1 | 2020-02-04 | 2020-02-04 | 0 |
| 1 | 2020-08-16 | 2021-03-16 | 1 |

##### Example 3

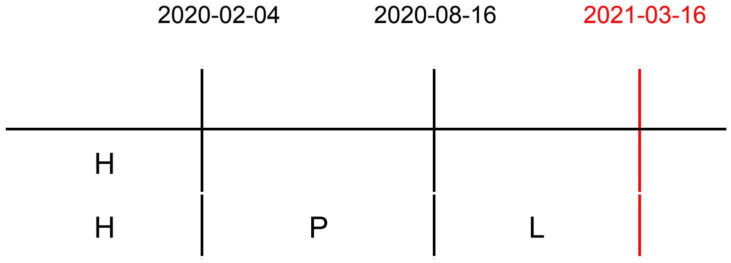

| Patient ID | Start date | Outcome date | label |
| --- | --- | --- | --- |
| 1 | 2020-02-04 | 2020-02-04 | 0 |
| 1 | 2020-08-16 | 2021-03-16 | 1 |
| 1 | 2021-08-09 | 2022-01-15 | 1 |

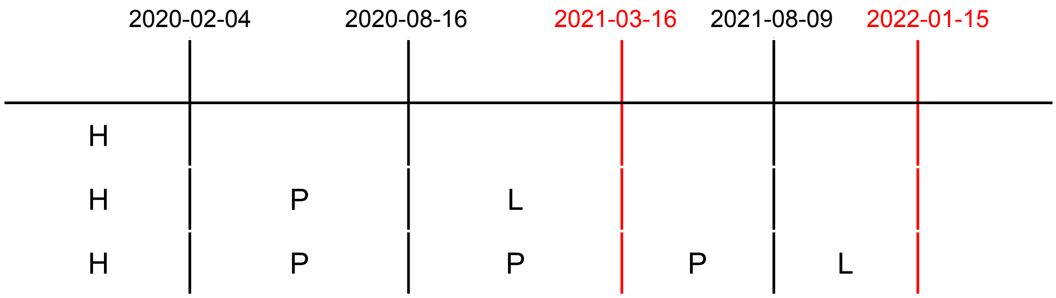


Scenario 4

| Patient ID | Start date | Outcome date | label |
| --- | --- | --- | --- |
| 1 | 2020-02-04 | 2020-02-04 | 0 |
| 1 | 2020-08-16 | 2021-03-16 | 1 |
| 1 | 2022-04-05 | 2022-04-05 | 0 |

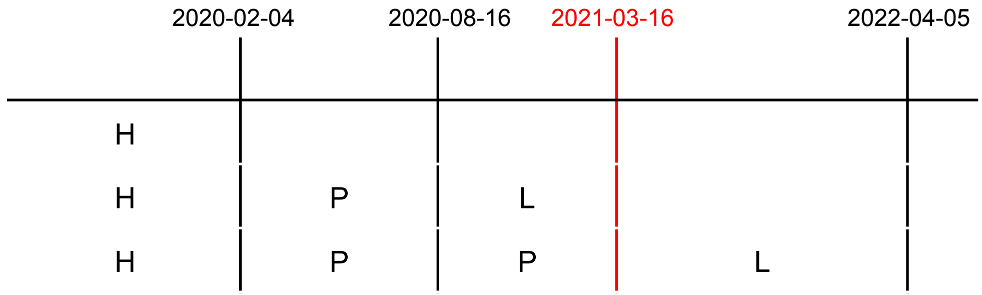


Example 5

| Patient ID | Start date | Outcome date | label |
| --- | --- | --- | --- |
| 1 | 2020-02-04 | 2020-02-04 | 0 |
| 1 | 2020-08-16 | 2020-08-16 | 0 |
| 1 | 2021-04-05 | 2021-10-05 | 1 |

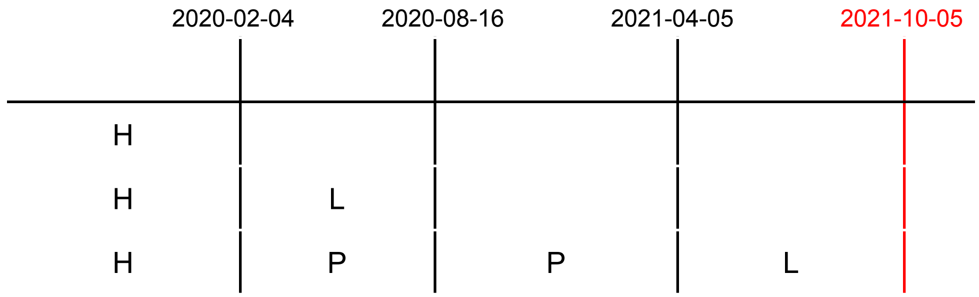


Example 6

| Patient ID | Start date | Outcome date | label |
|---|---|---|---|
| 1 | 2020-02-04 | 2020-02-04 | 0 |
| 1 | 2020-08-16 | 2020-08-16 | 0 |
| 1 | 2021-01-01 | 2021-01-01 | 0 |
| 1 | 2021-04-05 | 2021-10-05 | 1 |

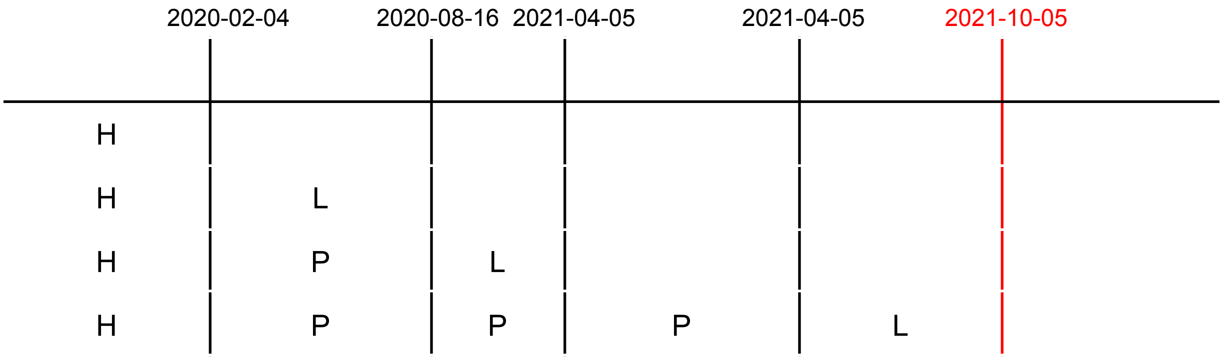


Example 7

| Patient ID | Start date | Outcome date | label |
|---|---|---|---|
| 1 | 2020-02-04 | 2020-02-04 | 0 |
| 1 | 2020-08-16 | 2021-03-16 | 1 |
| 1 | 2022-04-05 | 2022-04-05 | 0 |
| 1 | 2022-07-09 | 2023-02-04 | 1 |

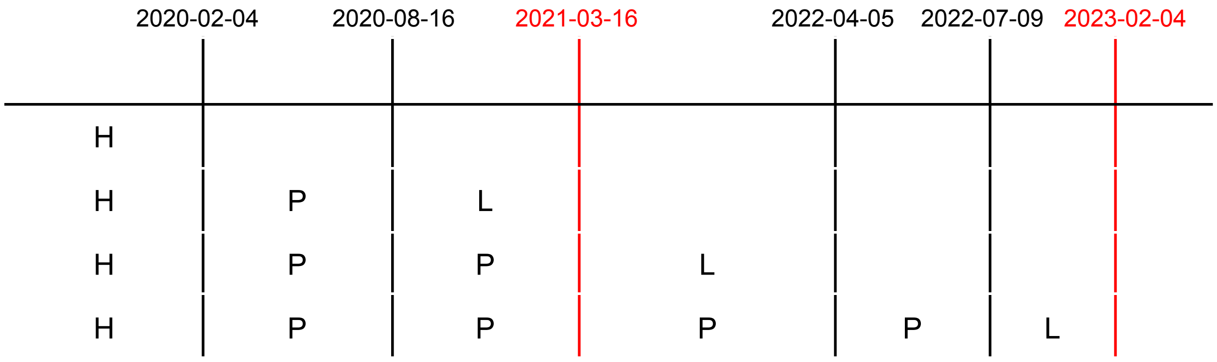
2020-02-04
2020-08-16
2021-03-16
2022-04-05
2022-07-09
2023-02-04
H
H P L
H P P L
H P P P P L
