## Supplemental Tables for "Temporal Learning with Dynamic Range (TLDR) for Modeling Recurrent Exposure and Treatment Outcomes"

**Table 1S**. Top features identified by the Gradient Boosting Model utilizing the TLDR approach

| Features | Description | Mean Feature Importance Score | Label | Rank |
|---|---|---|---|---|
| A17837157_past | Z20 Contact With And (Suspected) Exposure To Communicable Diseases | 96.815841 | past | 1 |
| A23575677_past | Blood Count | 64.010118 | past | 2 |
| A17864418_last | I10 Essential (Primary) Hypertension | 55.639180 | last | 3 |
| A23569177_past | Diagnostic Radiology (Diagnostic Imaging) Procedures Of The Chest | 53.816960 | past | 4 |
| A23574871_past | Cardiography Procedures | 47.795656 | past | 5 |
| A23569966_last | Surgical Procedures On Arteries And Veins | 42.956122 | last | 6 |
| A23569966_past | Surgical Procedures On Arteries And Veins | 36.223728 | past | 7 |
| A17864418_past | I10 Essential (Primary) Hypertension | 18.623674 | past | 8 |
| A23575677_last | Blood Count | 18.561787 | last | 9 |
| A17837257_last | Z79 Long Term (Current) Drug Therapy | 14.940546 | last | 10 |
| A17867143_past | R09 Other Symptoms And Signs Involving The Circulatory And Respiratory System | 10.178036 | past | 11 |
| A17825389_last | E11 Type 2 Diabetes Mellitus | 9.447193 | last | 12 |
| A17790457_past | R06 Abnormalities Of Breathing | 8.262680 | past | 13 |
| A17841649_history | R00 Abnormalities Of Heart Beat | 7.846064 | history | 14 |
| A17867137_past | R05 Cough | 7.266731 | past | 15 |
| A23574871_last | Cardiography Procedures | 6.070891 | last | 16 |
| A23569177_last | Diagnostic Radiology (Diagnostic Imaging) Procedures Of The Chest | 5.989005 | last | 17 |
| A17854422_history | R63 Symptoms And Signs Concerning Food And Fluid Intake | 5.345901 | history | 18 |
| A21035381_past | Basic Metabolic Panel This Panel Must Include The Following: Calcium (82310) Carbon Dioxide (82374) Chloride | 5.031301 | past | 19 |

| Features | Description | Mean Feature Importance Score | Label | Rank |
|---|---|---|---|---|
| | (82435) Creatinine (82565) Glucose (82947) Potassium (84132) Sodium (84295) Urea Nitrogen (Bun) (84520) (80048) | | | |
| A17812728_last | F41 Other Anxiety Disorders | 4.231843 | last | 20 |
| A17803345_history | R60 Edema, Not Elsewhere Classified | 3.547073 | history | 21 |
| A17799975_history | G47 Sleep Disorders | 3.285350 | history | 22 |
| LOINC:LP14355-9_past | Creatinine | 3.227448 | past | 23 |
| A17837257_past | Z79 Long Term (Current) Drug Therapy | 3.143967 | past | 24 |
| VANDF:CN103_last | Non-Opioid Analgesics (CN103) | 3.042145 | last | 25 |
| A17852637_history | M19 Other And Unspecified Osteoarthritis | 2.992135 | history | 26 |
| A17865919_history | M79 Other And Unspecified Soft Tissue Disorders, Not Elsewhere Classified | 2.981660 | history | 27 |
| A23575696_history | Echocardiography Procedures | 2.865306 | history | 28 |
| A17826701_history | K21 Gastro-Esophageal Reflux Disease | 2.855569 | history | 29 |
| A17837157_history | Z20 Contact With And (Suspected) Exposure To Communicable Diseases | 2.752461 | history | 30 |
| A17841684_history | R26 Abnormalities Of Gait And Mobility | 2.699457 | history | 31 |
| A17812728_history | F41 Other Anxiety Disorders | 2.567475 | history | 32 |
| A17803338_history | R52 Pain, Unspecified | 2.522947 | history | 33 |
| A17775558_history | K59 Other Functional Intestinal Disorders | 2.522372 | history | 34 |
| A17816202_past | R50 Fever Of Other And Unknown Origin | 2.513426 | past | 35 |
| LOINC:LP14635-4_past | Glucose | 2.471654 | past | 36 |
| A17787879_history | I49 Other Cardiac Arrhythmias | 2.307175 | history | 37 |
| A17790457_history | R06 Abnormalities Of Breathing | 2.165515 | history | 38 |
| A17849899_last | Z23 Encounter For Immunization | 2.145641 | last | 39 |

| Features | Description | Mean Feature Importance Score | Label | Rank |
|---|---|---|---|---|
| VANDF:TN102_past | IV Solutions With Electrolytes (TN102) | 2.093377 | past | 40 |
| A17838107_last | E66 Overweight And Obesity | 2.017368 | last | 41 |
| A17777747_history | R41 Other Symptoms And Signs Involving Cognitive Functions And Awareness | 1.981737 | history | 42 |
| VANDF:TN101_past | IV Solutions Without Electrolytes (TN101) | 1.962253 | past | 43 |
| A17777705_history | R19 Other Symptoms And Signs Involving The Digestive System And Abdomen | 1.943774 | history | 44 |
| A17852847_history | M47 Spondylosis | 1.942515 | history | 45 |

**Table 2S**. Top features identified by the Gradient Boosting Model utilizing the benchmark approach

| Features | Description | Mean Feature Importance Score | Rank |
|---|---|---|---|
| A17837157 | Z20 Contact With And (Suspected) Exposure To Communicable Diseases | 99.1786719 | 1 |
| A23569177 | Diagnostic Radiology (Diagnostic Imaging) Procedures Of The Chest | 37.4549334 | 2 |
| A17841649 | R00 Abnormalities Of Heart Beat | 25.7892871 | 3 |
| A17790457 | R06 Abnormalities Of Breathing | 24.1078640 | 4 |
| A17867143 | R09 Other Symptoms And Signs Involving The Circulatory And Respiratory System | 18.8931794 | 5 |
| A23574871 | Cardiography Procedures | 15.5573929 | 6 |
| A17803345 | R60 Edema, Not Elsewhere Classified | 15.2692797 | 7 |
| A23575696 | Echocardiography Procedures | 11.1512502 | 8 |
| A17854422 | R63 Symptoms And Signs Concerning Food And Fluid Intake | 10.8177669 | 9 |
| A20154187 | R91 Abnormal Findings On Diagnostic Imaging Of Lung | 10.1874953 | 10 |
| A1108251 | Troponin, Quantitative (84484) | 10.0626337 | 11 |
| A17812590 | E87 Other Disorders Of Fluid, Electrolyte And Acid-Base Balance | 9.0369827 | 12 |
| A17864418 | I10 Essential (Primary) Hypertension | 8.4008440 | 13 |
| A17865919 | M79 Other And Unspecified Soft Tissue Disorders, Not Elsewhere Classified | 8.2057906 | 14 |
| A17841684 | R26 Abnormalities Of Gait And Mobility | 7.9722797 | 15 |
| A2408688 | Natriuretic Peptide (83880) | 7.5507876 | 16 |
| A17790528 | R53 Malaise And Fatigue | 7.3122227 | 17 |
| A3858756 | Diagnostic Radiology Procedures | 7.2466181 | 18 |
| A17837257 | Z79 Long Term (Current) Drug Therapy | 7.2233035 | 19 |
| A17798771 | Z71 Persons Encountering Health Services For Other Counseling And Medical Advice, Not Elsewhere Classified | 6.6789242 | 20 |

| Features | Description | Mean Feature Importance Score | Rank |
|---|---|---|---|
| A17777747 | R41 Other Symptoms And Signs Involving Cognitive Functions And Awareness | 6.2493656 | 21 |
| A17853537 | N39 Other Disorders Of Urinary System | 6.2480706 | 22 |
| A20160123 | Z68 Body Mass Index (Bmi) | 6.2154824 | 23 |
| A17852637 | M19 Other And Unspecified Osteoarthritis | 6.1075750 | 24 |
| A17777705 | R19 Other Symptoms And Signs Involving The Digestive System And Abdomen | 5.9679031 | 25 |
| A23572454 | C-Reactive Protein | 5.6782729 | 26 |
| A17777752 | R42 Dizziness And Giddiness | 5.6626133 | 27 |
| A17825389 | E11 Type 2 Diabetes Mellitus | 5.3709465 | 28 |
| A17799975 | G47 Sleep Disorders | 5.1940295 | 29 |
| A23579008 | Prothrombin Time | 5.1400347 | 30 |
| VANDF:TN101 | IV Solutions Without Electrolytes (TN101) | 0.7240238 | 204 |

**Table 3S**. Examples of feature interpretation differences between two approaches

| Feature | TLDR | Benchmark |
|---|---|---|
| Z79 Long Term (Current) Drug Therapy | Last: #10 | #19 |
| Surgical Procedures On Arteries And Veins | Last: #6; Past: #7 | N/A |
| G47 Sleep Disorders | History: #22 | #29 |
| IV Solutions | Last: #40; Past: #43 | #204 |
